# Assessing the Impact of Area Deprivation Index on COVID-19 Prevalence: A Contrast Between Rural and Urban U.S. Jurisdictions

**DOI:** 10.1101/2020.10.07.20208462

**Authors:** Christopher Kitchen, Elham Hatef, Hsien-Yen Chang, Jonathan Weiner, Hadi Kharrazi

## Abstract

**Background:** The COVID-19 pandemic has impacted communities differentially, with poorer and minority populations being more adversely affected. Prior rural health research suggests such disparities may be exacerbated during the pandemic and in remote parts of the U.S.

**Objectives:** To understand the spread and impact of COVID-19 across the U.S., county level data for confirmed cases of COVID-19 were examined by Area Deprivation Index (ADI) scores and Metropolitan vs. Nonmetropolitan designations from the National Center for Health Statistics (NCHS). These designations were the basis for making comparisons between Urban and Rural jurisdictions.

**Methods:** Kendall’s Tau-B was used to compare effect sizes between jurisdictions on select ADI composites and well researched social determinants of health (SDH). Spearman coefficients and a moderation analysis using Poisson modeling was used to explore the relationship between ADI and COVID-19 prevalence in the context of county designation.

**Results:** Results show that the relationship between area deprivation and COVID-19 prevalence was positive and higher for rural counties, when compared to urban ones and that family income and poverty had a stronger relationship with prevalence than other ADI component measures.

**Conclusions:** Though most Americans live in Metropolitan Areas, rural communities were found to be associated with a stronger relationship between deprivation and COVID-19 prevalence. Models for predicting COVID-19 prevalence by ADI and county type reinforced this observation but revealed no moderating effect of county type on ADI.

## INTRODUCTION

The 2019-2020 coronavirus pandemic has underscored many of the public health disparities in the United States. Minority communities and people living in poverty account for disproportionately more COVID-19 cases and fatalities.^1,2^ The same communities may be inherently more vulnerable to infectious diseases due to underlying health conditions and lack of access to care.^3^ Past health disparities research has established a relationship between poor health outcomes and low socioeconomic status, often taken as a ranked measure of geographic area deprivation, or ADI.^4,5^ To date, few researchers have made use of ADI when evaluating COVID-19 prevalence across U.S. geographies, but early evidence seems to confirm a general positive relationship between deprivation and prevalence exists. ^6^ Proper disease management and policy efforts must understand these contrasts and public health needs to properly combat the spread of COVID-19. ^7^ ADI is an important tool for this discovery.

To date, less attention has been given to the spread of COVID-19 in rural communities, even though recent evidence suggests a rapid spread in rural areas.^8^ Greater prevalence of chronic disease and remoteness of these areas are cause for concern, even though they make up only a fraction of COVID-19 cases in the U.S. ^9,10,8^ Rural communities also tend to have worse prospects for healthcare access and outcomes.^11- 14^ By extension, we may expect poorer outcomes for more impoverished rural jurisdictions during the pandemic.^13,14^

ADI is used in this analysis as a predictor for COVID-19 prevalence that permits contrast between diverse communities. We classified 3,142 counties across the U.S. as “Urban” or “Rural” and stratified the relationship between prevalence and ADI accordingly. Our hypothesis is that ADI is predictive of COVID-19 prevalence and moderated by county type.

## METHODS

### Data Sources

Current estimates for COVID-19 cases were obtained from the JHU CSSE Coronavirus tracking project.^15,16^ This data repository contains county level time series data for confirmed cases dating back to January 22^nd^, 2020 and commonly used by population health researchers for modeling COVID-19 spread.^17,18,19^ Estimates for August 20^th^, 2020 were used for each of the 3142 US counties for this analysis. Population by race/ethnicity, and gender per county were based on 2019 estimates from the 2010 U.S. Census.^20,21^ Case prevalence was calculated as a count of confirmed cases per 100k persons in each county.

### Area Deprivation Index

County level ADI is constructed from weighting 17 widely used measures in population health literature for poverty, income, and education.^4,22^ The 5-year estimates of 2018 American Community Survey (ACS) data were used for calculating ADI and each of the composite measures, using an approach as described by Singh et al. ^4,21^ Higher raw ADI corresponds to more deprivation and therefore lower socioeconomic status (SES). A low ADI national percentile rank corresponds to high raw ADI and more deprivation, or in other words the 100^th^ percentile rank has the most disadvantaged counties at a national level.

### Urban vs Rural Designation

The National Center for Health Statistics (NCHS) first developed a mechanism for classifying rural and urbanized areas at the county level in 2001 for the accurate assessment and measurement of health differences between residents.^23,24,25^ The 2013 NCHS Urbanization scheme defines Metropolitan Statistical Areas (MSA) as at least 50,000 residents with an urban nucleus of at least 1,000 persons per square mile. Counties that contain an urbanized core are considered central counties for the purpose of defining the MSA. Neighboring counties with a density of at least 500 are also included in the MSA designation. Nonmetropolitan counties (hereafter, “Rural”) are micropolitan or noncore geographies of fewer than 50,000 residents. 2013 NCHS definitions for metropolitan and nonmetropolitan jurisdictions were collapsed to a binary variable reflecting Urban and Rural county type.

### Statistical Tests

Descriptive characteristics for population, population density, ADI, select ADI components, Census variables and CSSE COVID-19 case figures were tabulated across county type. Effect sizes for each characteristic by binary county type were estimated using Kendall’s tau (positive effect corresponds to “Urban” county type). Additional county-level social determinants of health (SDH) variables included percent male, percent non-Caucasian minority and percent aged 65 years or older. A subset of SDH variables are presented in this work to reduce redundancy of ADI measures, while illustrating resident demographics and several relevant domains of the ADI.

Spearman correlation coefficients were calculated for a subset of relevant county characteristics, ADI variables and CSSE COVID-19 estimates and for each county type. These included median family income, percent living under 150% of poverty percent of households without a vehicle, and percent of households with more than one person per bedroom.

Finally, three models using logarithmic link functions were fitted to explore a moderation effect of county type (i.e. “Urban”, vs. “Rural”) and ADI on COVID-19 case prevalence. A base comparison model is defined as a mapping of ADI national rank to prevalence. The first test model fitted ADI and county type separately, and the second was the same but with an interaction term. Inspection of coefficient sizes allowed us to interpret whether county type had a moderating effect on ADI.

## RESULTS

### Characteristics of Urban vs. Rural

Table 1 reflects common SDH, including household income (in USD), percent of families below poverty, percent of households with no vehicles and percent of households with more than one person per room. Rural counties were found to have significantly worse outcomes, including median family income (mean=$59,097), percent of residents under 150% of poverty (mean=12%), and characteristically more male (mean=50.4%), more non-Caucasian residents (mean=15.4%) and residents aged 65 or older (mean=17.1%). No significant difference was found in percent of households with more than one occupant per room (mean=2.5). Rural counties had significantly fewer COVID-19 cases, cases per capita and deaths as of August 20, 2020 (mean=290.33; 1,172.4; 6.6).

**Table 1.** Population characteristics for ADI, SDH variables, and COVID-19 case prevalence between Rural and Urban county types.

| Feature | Rural | Urban | Effect Size ($\tau$) | All Counties |
| --- | --- | --- | --- | --- |
| **Count of Counties** | 1976 (62.9%) | 1166 (37.1%) | - | 3142 (100%) |
| **Characteristics** |  |  |  |  |
| **Total Population** | 46063061 (13.8%) | 288884573 (86.2%) | - | 334947634 (100%) |
| **Density** | 42.9 (95) | 625.7 (2790.7) | 0.493 ($p<.001$) | 259.3 (1724.9) |
| **ADI Variables** |  |  |  |  |
| **Raw ADI** | 103.1 (15.9) | 90.5 (17.1) | -0.274 ($p<.001$) | 98.4 (17.4) |
| **National Rank ADI** | 56.6 (26.5) | 37 (26.7) | -0.276 ($p<.001$) | 49.3 (28.2) |
| **Median Family Income (USD)** | 59097.3 (12051.8) | 72494.4 (17863.5) | 0.323 ($p<.001$) | 64069 (15861.5) |
| **% Families in Poverty** | 12 (6.1) | 9.9 (4.4) | -0.136 ($p<.001$) | 11.2 (5.7) |
| **% No Vehicle Households** | 6.4 (4.5) | 6.3 (4.5) | -0.034 ($p=.021$) | 6.4 (4.5) |
| **% > 1 Person per Room Households** | 2.5 (2.8) | 2.4 (1.8) | 0.019 ($p=.184$) | 2.4 (2.4) |
| **Other SDH** |  |  |  |  |
| **% Male** | 50.4 (2.7) | 49.5 (1.7) | -0.184 ($p<.001$) | 50.1 (2.4) |
| **% Non-Caucasian Race** | 15.4 (17.5) | 19.6 (15.4) | 0.189 ($p<.001$) | 16.9 (16.9) |
| **% 65yoa or Older** | 17.1 (3.9) | 14.4 (3.5) | -0.296 ($p<.001$) | 16.1 (4) |
| **COVID-19 Case-Prevalence Info** |  |  |  |  |
| **Confirmed Cases** | 290.3 (439.7) | 4170.4 (13483.7) | 0.435 ($p<.001$) | 1730.2 (8430.3) |
| **Cases per Capita** | 1172.4 (1248.7) | 1362.7 (1063.9) | 0.156 ($p<.001$) | 1243 (1186.9) |
| **Deaths** | 6.6 (14) | 135.1 (771.9) | 0.41 ($p<.001$) | 54.3 (474.3) |

**Table 2.**
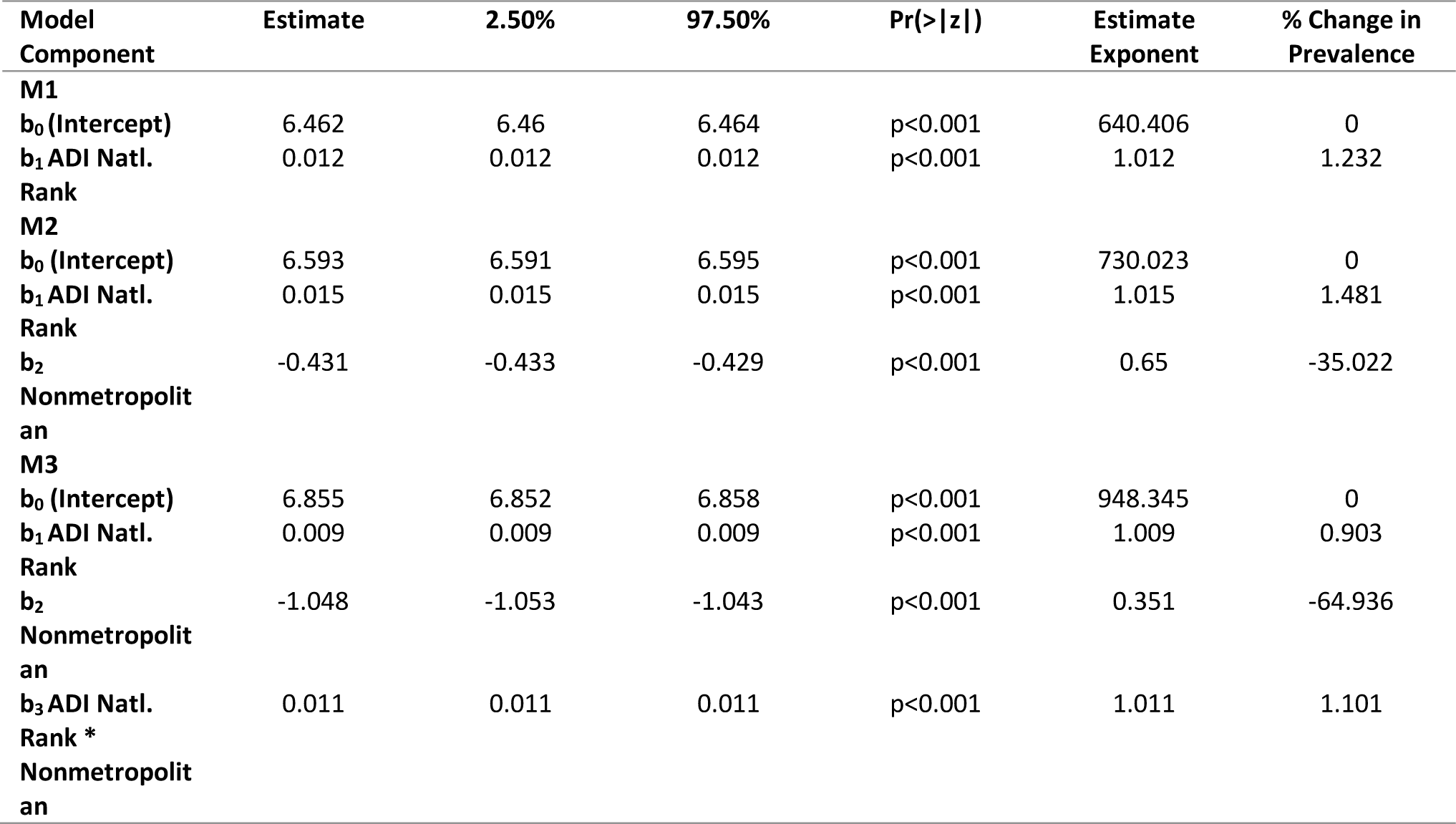
Model performance and coefficients obtained in ADI vs County Type moderation analysis.

### Correlation between Prevalence and ADI by County Type

COVID-19 prevalence is higher in Urban counties, but less correlated to national ADI rank when compared to Rural (ρ=.25;. 46, respectively) (Figure 1). Prevalence for Urban counties was also less strongly correlated with family income (ρ=-.17; -.32), percent of households under 150% of poverty (ρ=.29; .39), and without a vehicle (ρ=.16; .21) when compared to Rural counties. Similar coefficients between Urban and Rural were found for prevalence and percent of households with more than one person per room (ρ=.37; .33).

**Figure 1.**
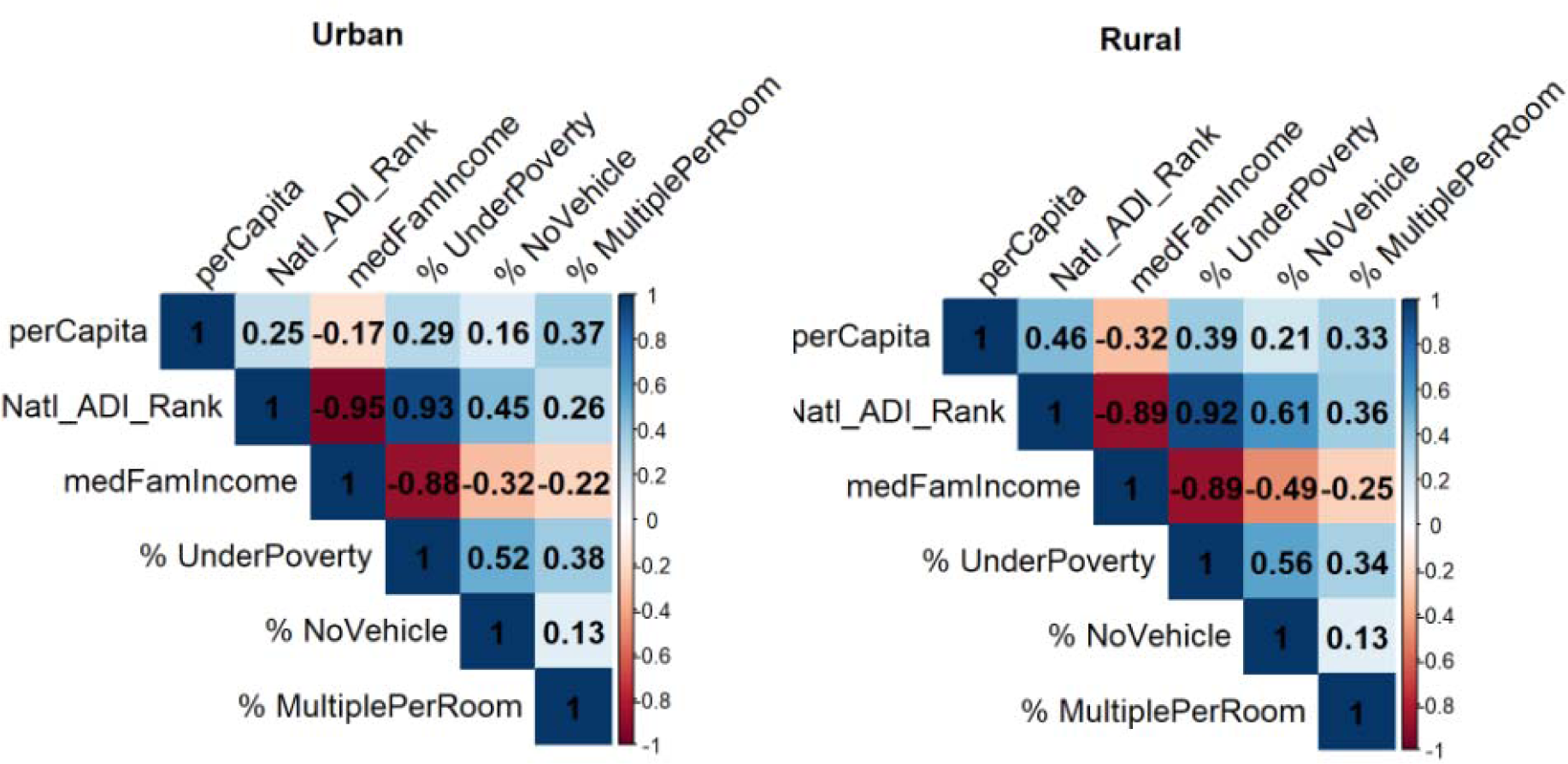
Correlation matrices for select COVID-19 case prevalence and ADI components across Urban and Rural county types.

### Modeling Prevalence by ADI and County Type

The base model for prevalence as a function of ADI yielded a significant parameter estimate (p<.001), but only accounted for around 1.2% change in prevalence and had rather poor overall fit (AIC=2378220). ADI with county type improved overall model prediction by about 6.4% (AIC=2226173), largely stemming from a 35% decrease in prevalence due to county type. The inclusion of the interaction term in the second test model also improved model fit over base by 9.6% (AIC=2150645). The county type parameter grew, accounting for 65% of change, while ADI within Urban and Rural counties fell to around 1% (0.9% and 1.1%, respectively).

## DISCUSSION

Although discrepancies between Urban and Rural county jurisdictions were evident in SDH measures, no substantial moderating effect could be discerned using Poisson regression modeling. The coefficients obtained for ADI within Urban and Rural jurisdictions were very similar but larger for Rural jurisdictions. Each was smaller than the coefficient obtained in the base model for ADI, revealing no moderation effect.

Model fit was also consistently poor, likely due to the high variance in COVID-19 cases across counties. Several ADI components, however, reached levels of significance for Spearman correlation with COVID-19 prevalence. Among these, Urban jurisdictions tended to have lower correlation with COVID-19 prevalence than Rural ones. Together, these results suggest that (1) the overall prevalence of COVID-19 is lower among rural jurisdictions, and (2) the effect of socioeconomic disparity on COVID-19 prevalence is worse for rural jurisdictions over urban ones.

These results require several qualifications. First, these results are a snapshot for an ongoing pandemic. The relationship between ADI, its component measures and COVID-19 prevalence are a fluid and changing phenomenon. Second, the granularity of both the classification scheme and level of geography are less than ideal for detecting small or more nuanced effects. We expect much greater heterogeneity in ADI composites for densely populated regions. Zip code, census tract or block group level data may have been more appropriate, but this information for COVID-19 testing results is not currently available nationwide.^26^ Third, ADI only captures a handful of SDH that, while useful and widely used, do not account for the very real possibility of racial disparity in COVID-19 spread. Race, age and gender should be considered in future modeling efforts for coronavirus prevalence. Finally, we only explored confirmed cases of coronavirus as an outcome measure, but there is at least as much justification for modeling case fatality. At present, little is known about the prevalence of asymptomatic cases, so the true impact of severe COVID-19 infection differential to county type may be even greater than what can currently be estimated.

Additional work is required to tie in known risk factors and SDH to adequately address long-standing disparities in health outcomes and predict geographies that are most impacted by a pandemic.^27^ Rural communities have notably different challenges to access care than those in more densely populated areas.^28,29,8^ During a pandemic, lack of reliable internet access and transportation may compound the effect of poverty on telehealth services or ambulatory care. Measures of cost or healthcare utilization could serve as proxy outcomes for severity of pandemic impact. Finally, as more data become available on coronavirus cases, we expect finer resolution of geographic data, making it necessary to reevaluate and confirm these findings in smaller community levels.

## Data Availability

Data used in the development of this manuscript were derived from publicly available files through the JHU CSSE Coronavirus tracking project and The U.S. Census Bureau's ACS.

## Acknowledgements

We thank our colleagues at the JHU Center for Systems Science and Engineering for managing a world class resource on COVID-19 data and the reviewers for their timely feedback on this manuscript during a time of public health crisis.

## Notes

Disclosures: No award or payment was given to any party involved in the production of this work. Each listed author made substantial contributions to the development of and analysis in this manuscript. The listed authors have no real or perceived conflicts of interest to disclose in the dissemination of this work.

### Competing Interest Statement

The authors have declared no competing interest.

### Funding Statement

No award or payment was given to any party involved in the production of this work.

### Author Declarations

This analysis makes use of publicly available data and was exempt from IRB review at Johns Hopkins University.

